# Better Antimicrobial Resistance Data Analysis & Reporting in Less Time

**DOI:** 10.1101/2021.07.06.21257599

**Authors:** Christian F. Luz, Matthijs S. Berends, Xuewei Zhou, Mariëtte Lokate, Alex W. Friedrich, Bhanu Sinha, Corinna Glasner

## Abstract

**Introduction:** The global challenge of antimicrobial resistances (AMR) requires the rational and responsible use of antimicrobials. Insights and knowledge about the local AMR levels and epidemiology are essential to guide optimal decision-making processes in antimicrobial use. However, dedicated tools for reliable and reproducible AMR data analysis and reporting are often lacking. Previously, we have developed a novel approach to AMR data analysis and reporting using open-source software tools. In this study, we aimed at comparing the effectiveness and efficiency of traditional analysis and reporting versus this new approach for reliable and reproducible AMR data analysis in a clinical setting.

**Methods:** Ten professionals in the field of AMR that routinely work with AMR data were recruited to participate and provided with one year’s blood culture test results from a tertiary care hospital results including antimicrobial susceptibility test results. Participants were asked to perform a detailed AMR data analysis in a two-step process: first (round 1) using their analysis software of choice and next (round 2) using the previously developed open-source software tools. Accuracy of the results and time spent were compared between the two rounds. Paired student’s t-tests were used to test for statistical significance. Finally, participants rated the usability of the tools using the systems usability scale.

**Results:** The mean time spent on creating a comprehensive AMR report reduced from 93.7 (SD ±21.6) minutes to 22.4 (SD ±13.7) minutes (p < 0.001). Average task completion per round changed from 56% (SD: ±23%) to 96% (SD: ±5.5%) (p<0.05). The proportion of correct answers in the available results increased from 37.9% in the first round to 97.9% in the second round (p < 0.001). The usability of the new AMR reporting tool was rated with a median of 83.8 (out of 100) on the system usability scale.

**Conclusion:** This study demonstrated the significant improvement in efficiency and accuracy in standard AMR data analysis and reporting workflows through the use of open-source software tools in a clinical setting. Integrating these tools in clinical settings can democratise the access to fast and reliable insights about local microbial epidemiology and associated AMR levels. Thereby, our approach can support evidence-based decision-making processes in the use of antimicrobials.

## Introduction

Antimicrobial resistance (AMR) is a global challenge in healthcare, livestock and agriculture, and the environment alike. The *silent tsunami* of AMR is already impacting our lives and the wave is constantly growing [1,2]. One crucial action point in the fight against AMR is the appropriate use of antimicrobials. The choice and use of antimicrobials has to be integrated into a well-informed decision making process and supported by antimicrobial and diagnostic stewardship programmes [3,4]. Next to essential local, national, and international guidelines on appropriate antimicrobial use, the information on AMR rates and antimicrobial use through reliable data analysis and reporting is vital. While data on national and international levels are typically easy to access through official reports, local data insights are often lacking, difficult to establish, and its generation requires highly trained professionals. Unfortunately, working with local AMR data is often furthermore complicated by very heterogeneous data structures and information systems within and between different settings [5,6]. Yet, decision makers in the clinical context need to be able to access these important data in an easy and rapid manner. Without a dedicated team of epidemiologically trained professionals, providing these insights could be challenging and error-prone. Incorrect data or data analyses could even lead to biased/erroneous empirical antimicrobial treatment policies.

To overcome these hurdles, we previously developed new approaches to AMR data analysis and reporting to empower any expert on any level working with or relying on AMR data [7,8]. We aimed at reliable, reproducible, and transparent AMR data analysis. The underlying concepts are based on open-source software, making them free to use and adaptable to any setting-specific needs. To specify, we developed a software package for the statistical language R to simplify and standardise AMR data analysis based on international guidelines [7]. In addition, we demonstrated the application of this software package to create interactive analysis tools for rapid and user-friendly AMR data analysis and reporting [8].

However, while the use of our approach in research has been demonstrated [9–12], the impact on workflows for AMR data analysis and reporting in clinical settings is pending. AMR data analysis and reporting are typically performed at clinical microbiology departments in hospitals, in microbiological laboratories, or as part of multidisciplinary antimicrobial stewardship activities. AMR data analysis and reporting require highly skilled professionals. In addition, thorough and in-depth analyses can be time consuming and sufficient resources need to be allocated for consistent and repeated reporting. This is further complicated by the lack of available software tools that fulfill all requirements such as incorporation of (inter-) national guidelines or reliable reference data.

In this study, we aimed at demonstrating and studying the usability of our developed approach and its impact on clinicians’ workflows in an institutional healthcare setting. The approach should enable better AMR data analysis and reporting in less time.

## Methods

The study was initiated at the University Medical Center Groningen (UMCG), a 1339-bed tertiary care hospital in the Northern Netherlands and performed across the UMCG and Certe (a regional laboratory) in the Northern Netherlands. It was designed as a comparison study to evaluate the efficiency, effectiveness, and usability of a new AMR data analysis and reporting approach [7,8] against traditional reporting.

### Study setup

The setup of the study is visualised in Figure 1 and is explained in the following sections.

**Figure 1.**
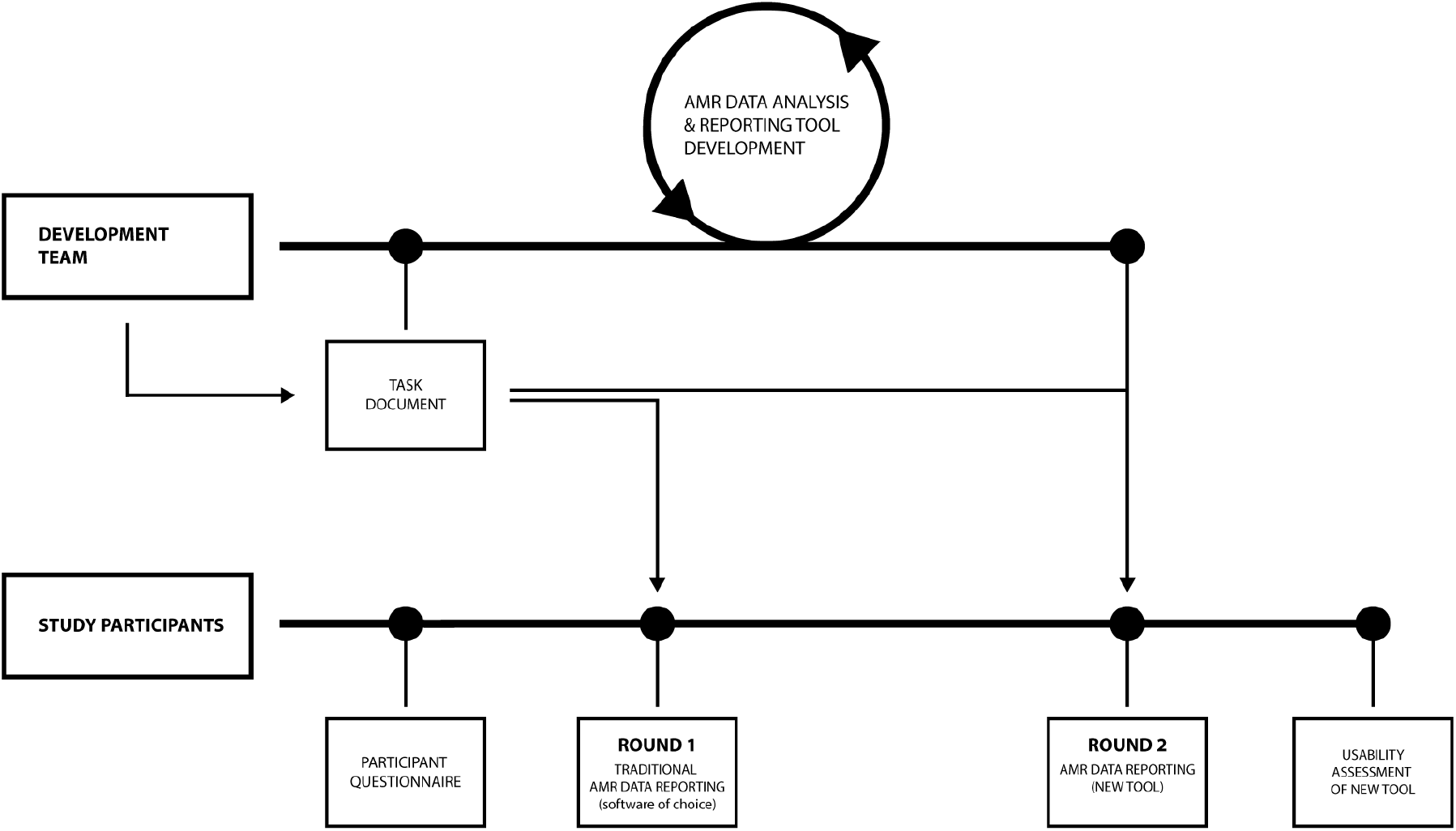
Study setup; the same AMR data was used along all steps and rounds.

The study was based on a task document listing general AMR data analysis and reporting tasks (Table 1). This list served as the basis to compare effectiveness (solvability of each task for every user) and efficiency (time spent solving each task) of both approaches. Tasks were grouped into five related groups and analyses were performed per group (further referred to as five tasks). A maximum amount of time per task (group) was defined for each task. The list of tasks including correct results is available in the appendix (A1)

**Table 1.** AMR data analysis and reporting tasks

| Task | Task description | Maximum time (minutes) |
| --- | --- | --- |
| 1 | Total number of blood culture sets per year | 15 |
| 2a | Total number of positive blood culture sets per year | 20 |
| 2b | Total number of negative blood culture sets per year |  |
| 3 | Top ten isolated microorganisms in blood cultures per year including isolate count (first isolates*) | 20 |
| 4a | Resistance profile (S/I & R) in *Escherichia coli* (first isolates*) found in blood cultures for selected antimicrobials |  |
| 4b | Resistance profile (S/I & R) in *Klebsiella pneumoniae* (first isolates*) found in blood cultures for selected antimicrobials | 30 |
| 4c | Resistance profile (S/I & R) in *Staphylococcus aureus* (first isolates*) found in blood cultures for selected antimicrobials |  |
| 5a | Empiric susceptibility rate for *Escherichia coli* and *Klebsiella pneumoniae* (first isolates* only for both) found in blood cultures with a combination of cefuroxime and tobramycin |  |
| 5b | Empiric susceptibility rate for *Escherichia coli* and *Klebsiella pneumoniae* (first isolates* only for both) found in blood cultures with a combination of amoxicillin & clavulanic acid and tobramycin OR amoxicillin & clavulanic acid and gentamicin | 30 |
| 5c | Empiric susceptibility rate for *Escherichia coli* and *Klebsiella pneumoniae* (first isolates* only for both) found in blood cultures with a combination of ceftriaxone and tobramycin OR ceftriaxone and gentamicin |  |

S = susceptible; I = susceptible, increased exposure; R = resistant \*) [13]

### AMR data

Anonymised microbiological data were obtained from the Department of Medical Microbiology and Infection Prevention at the UMCG. The data consisted of 23,416 records from 18,508 unique blood culture tests that were taken between January 1, 2019 and December 31, 2019 which were retrieved from the local laboratory information system (LIS). Data were collected retrospectively and permission was granted by the local ethical committee (METc 2014/530). Available variables were: test date, sample identification number, sample specimen, anonymised patient identification number, microbial identification code (if culture positive), antimicrobial susceptibility test results (S, I, R - susceptible, susceptible at increased exposure, resistant) for 52 antimicrobials. The exemplified data structure is presented in Table 2.

**Table 2.**
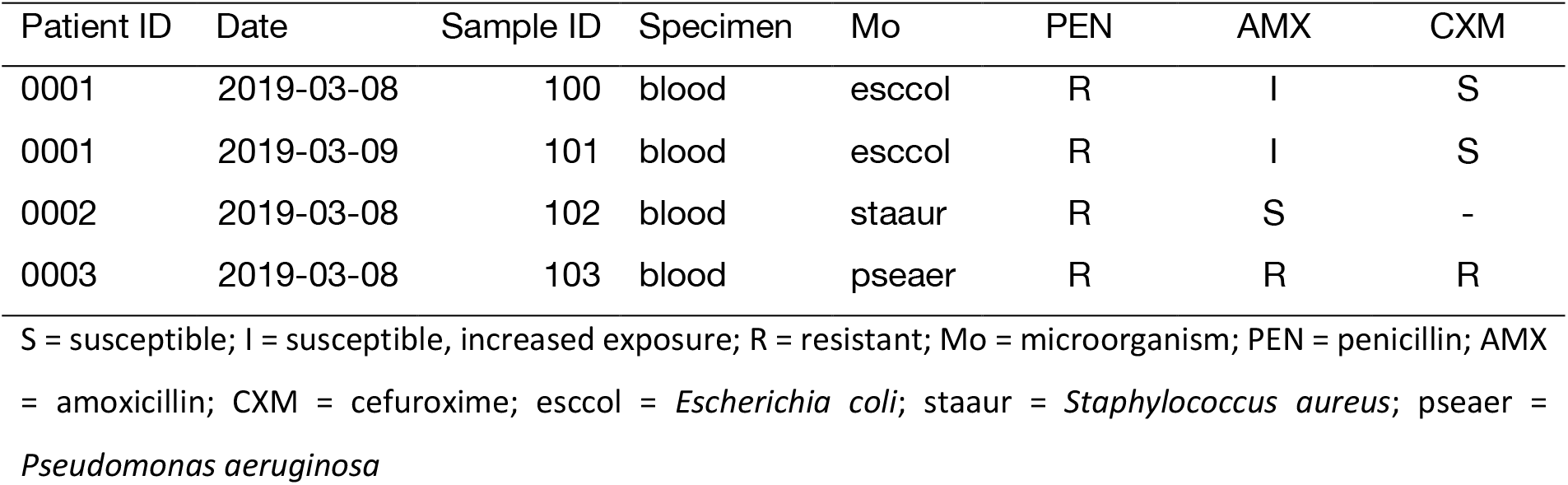
Raw data example

### AMR data analysis and reporting

.We used our previously developed approach [7,8] to create a customised browser-based AMR data analysis and reporting application. This application was used in this study and applied to the AMR data analysis and reporting tasks listed in the task document (Table 1). The development of the application followed an agile approach using scrum methodologies [14]. Agile development was used to effectively and iteratively work in a team of two developers (CFL, MSB), a clinical microbiologist (XZ), and an infection preventionist (ML). The application was designed as an interactive web-browser based dashboard (Figure 2). The prepared dataset was already loaded into the system and interaction with the application was possible through any web-browser.

**Figure 2.**
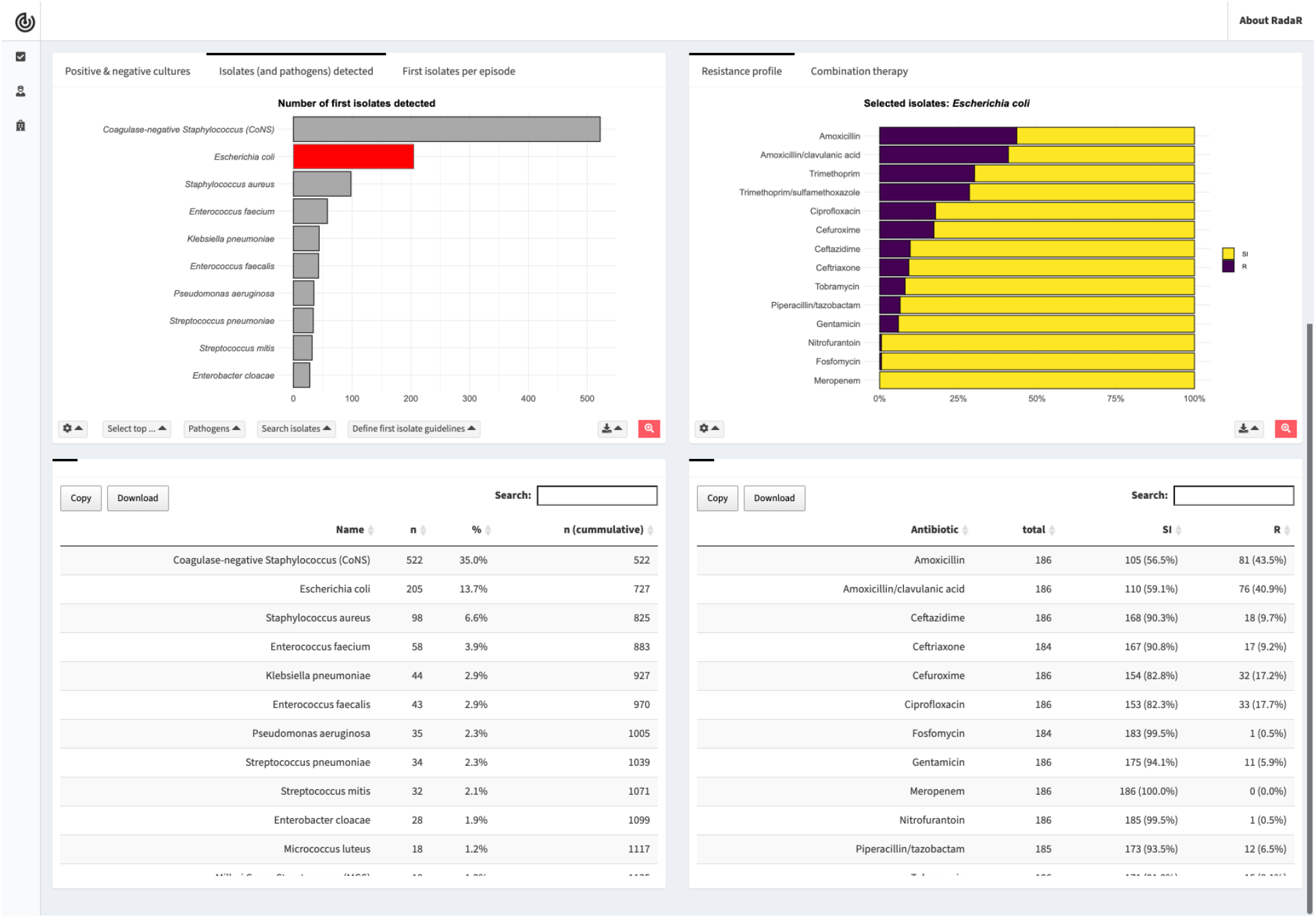
Interactive dashboard for AMR data analysis used in this study.

### Study participants

Participants in this study were recruited from the departments of Medical Microbiology, Critical Care Medicine, and Paediatrics, to reflect heterogeneous backgrounds of healthcare professionals working with AMR data. Members of the development team did not take part in the study.

### Study execution and data

First, study participants were asked to fill in an online questionnaire capturing their personal backgrounds, demographics, software experience, and experience in AMR data analysis and reporting. Next, participants were provided the task document together with the AMR data (csv- or xlsx-format). The participants were asked to perform a comprehensive AMR data report following the task document using their software of choice (round 1). Task results and information on time spent per task were self-monitored and returned by the participant using a structured report form. Lastly, participants repeated the AMR data analysis and reporting process with the same task document but using the new AMR data analysis and reporting application (round 2). Task results and information on time spent per task were again self-monitored and returned by the participant using the same structured report form as in the first round. This last step was evaluated using a second online questionnaire. The study execution process is illustrated in Figure 1.

### Evaluation and study data analysis

The utility of the new AMR data analysis and reporting application was evaluated according to ISO 9241-11:2018 [15]. This international standard comprises several specific metrics to quantify the usability of a tool with regard to reaching its defined goals (Figure 3). In this study the goal was a comprehensive AMR data report and comprised several tasks as outlined in the task document. The equipment was the focus of this study (traditional AMR data analysis and reporting approach vs. newly developed AMR data analysis and reporting approach).

**Figure 3.**
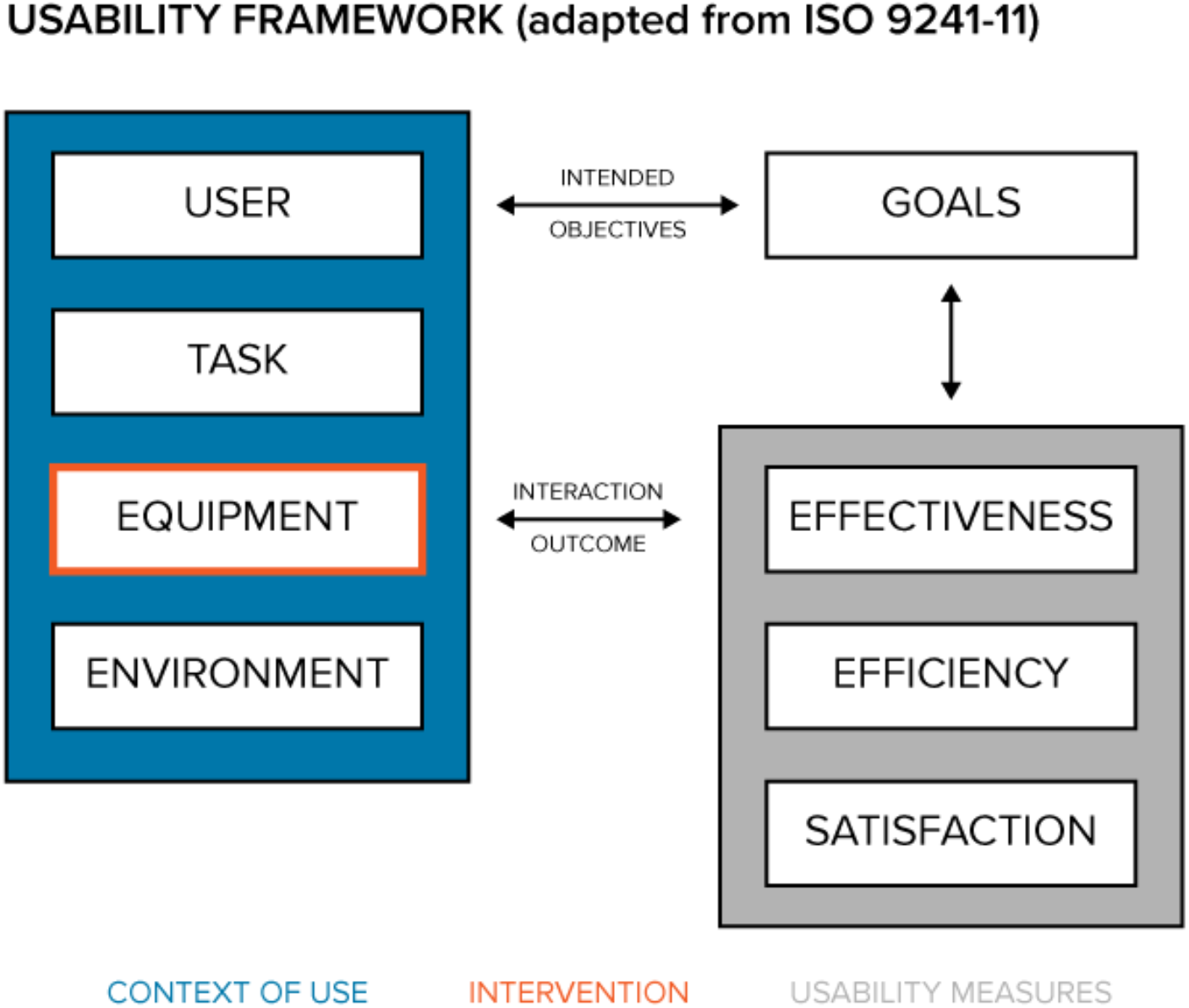
Usability framework based on ISO 9241-11

The three ISO standard usability measures (in grey) were defined as follows in this study: Effectiveness was determined by degree of task completion coded using three categories: 1) completed; 2) not completed (task not possible to complete); 3) not completed (task completion would take too long, e.g. > 20 minutes). In addition, effectiveness was assessed by the variance in the task results stratified by study round. Deviation from the correct results was measured in absolute percent from the correct result. To account for potential differences in the results due to rounding, all numeric results were transformed to integers. Efficiency was determined by timing each individual task. Time on task started when the user started performing the task, all data was loaded, and the chosen analysis software was up and running. Time on task ended when the task reached one of the endpoints, as described above. In the analysis, the mean time for each task and the mean total time for the complete report across users was calculated. Statistical significant difference was tested using paired Student’s t-test. All analyses were performed in R [16]. Outcomes of tests were considered statistically significant for *p* < 0.05. Accuray of the reported results per task and round were studied by calculating the deviation of the reported result in absolute percent from the correct result. Satisfaction was measured using the System Usability Scale (SUS), a 10-item Likert scale with levels from 1 (strongly disagree) to 5 (strongly agree, see Appendix 3) [17]. The SUS yields a single number from 0 to 100 representing a composite measure of the overall usability of the system being studied (SUS questions and score calculation in the Appendix A2).

## Results

### Study participants

In total 10 participants were recruited for this study. Most participants were clinical microbiologists (in training) (70%). The median age of the participant group was 40.5 years with a median working experience in the field of 8.0 years. The relevance of AMR data as part of the participants’ job was rated very high (median of 5.0; scale 1-5). AMR data analysis was part of the participants’ job for 60% of all participants. Participants reported to be very experienced in interpreting AMR data structures (median 5.0, scale 1-5). Participants were less experienced in epidemiological data analysis (median 3.0, scale 1-5). All participant characteristics are summarised in Table 3.

**Table 3.**
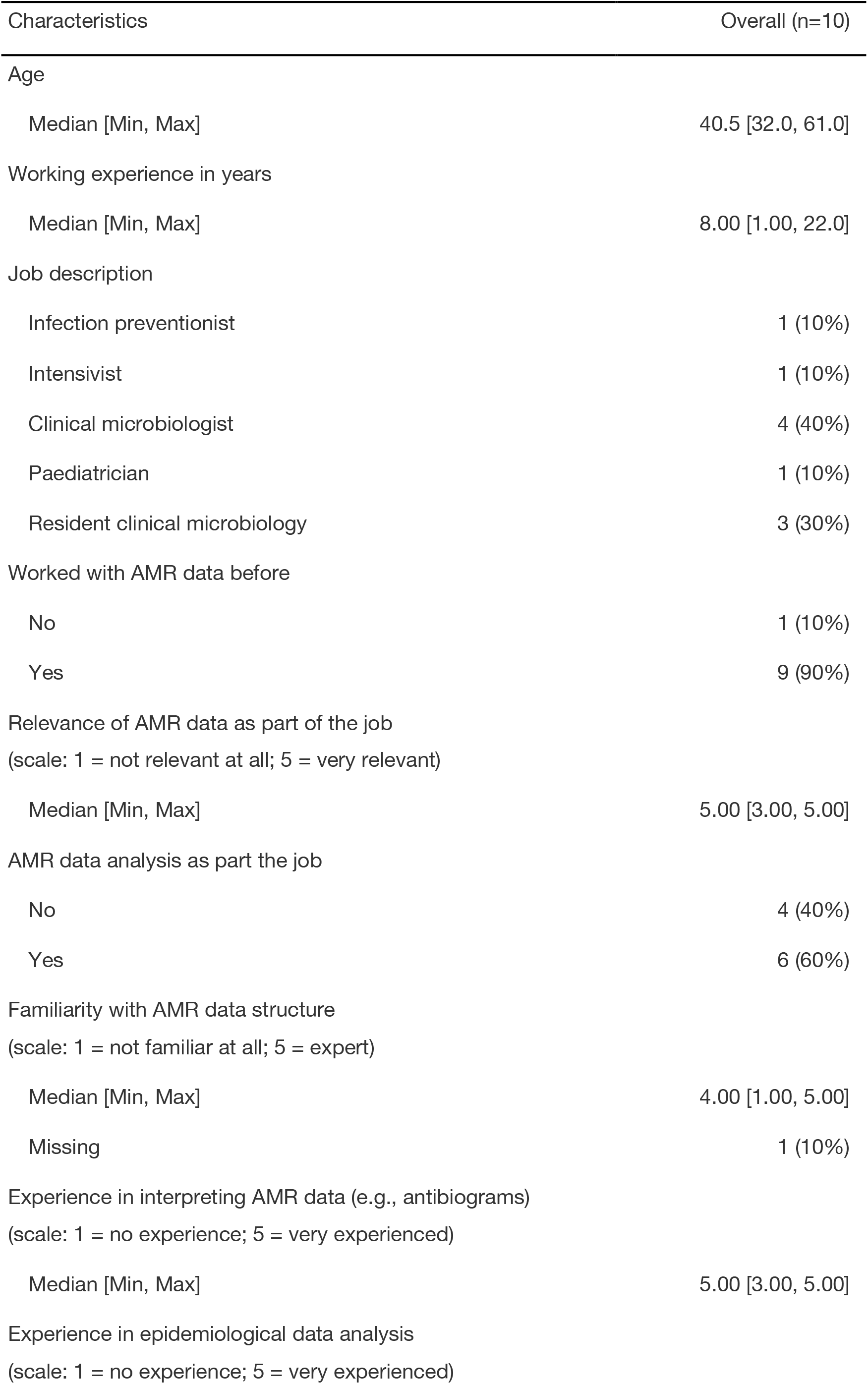
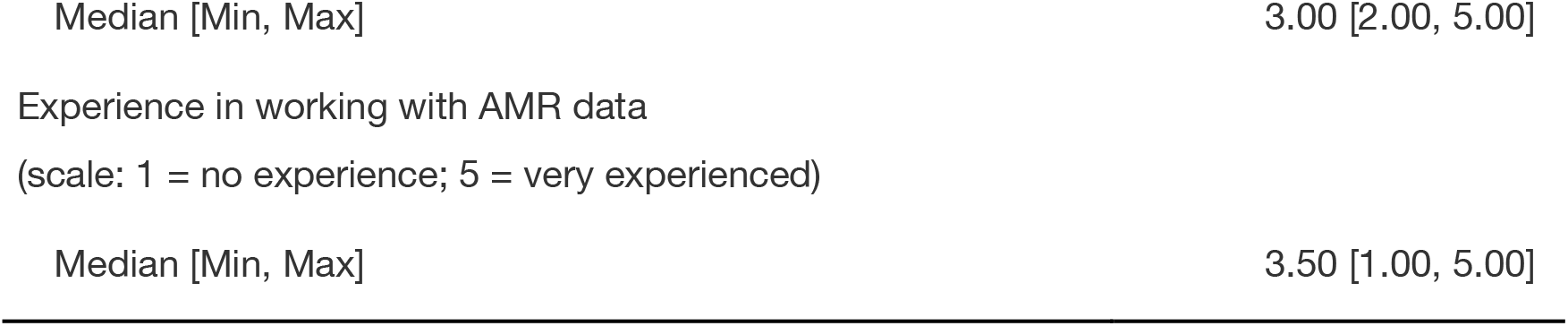
Study participant characteristics

The participants reported a diverse background in software experience for data analysis, with most experience reported for Microsoft Excel (Figure 4).

**Figure 4.**
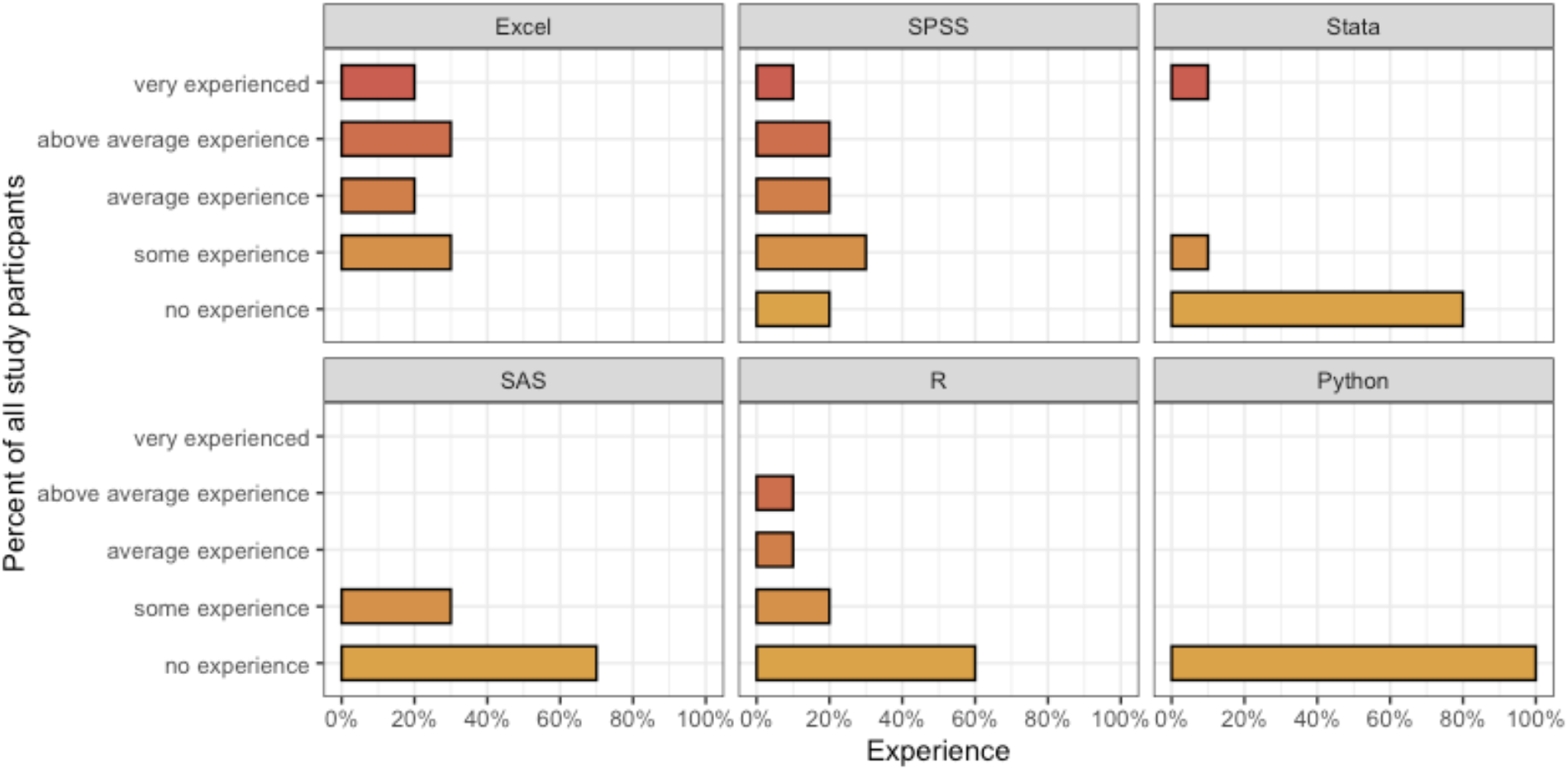
Data analysis software experience reported by study participants.

### Effectiveness and accuracy

Not all participants were able to complete the tasks within the given time frame. Average task completion between the first round (traditional AMR data analysis and reporting) and the second round (new AMR data analysis and reporting) changed from 56% (SD: 23%) to 96% (SD: 6%) (p < 0.05). Task completion per question and round is displayed in Figure 5. Variation in responses for each given task showed significant differences between the first and second round. Figure 6 shows the deviation in absolute percent from the correct results from the correct result per round and task. The proportion of correct answers in the available results increased from 38% in the first round to 98% in the second round (p < 0.001). A sub-analysis of species-specific results for task 3 round 1 is available in the appendix (A3).

**Figure 5.**
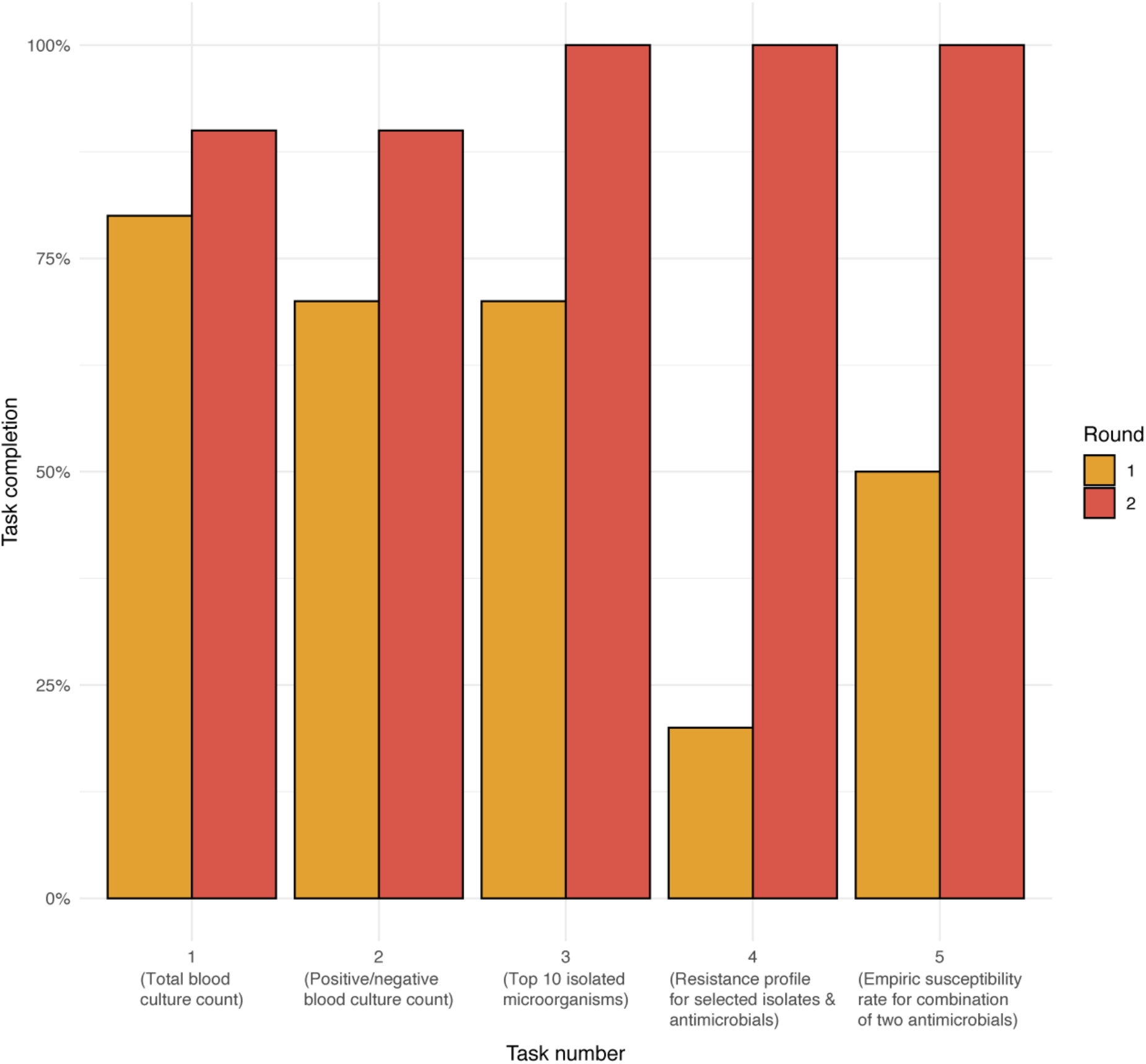
Task completion in percent by task number and round.

**Figure 6.**
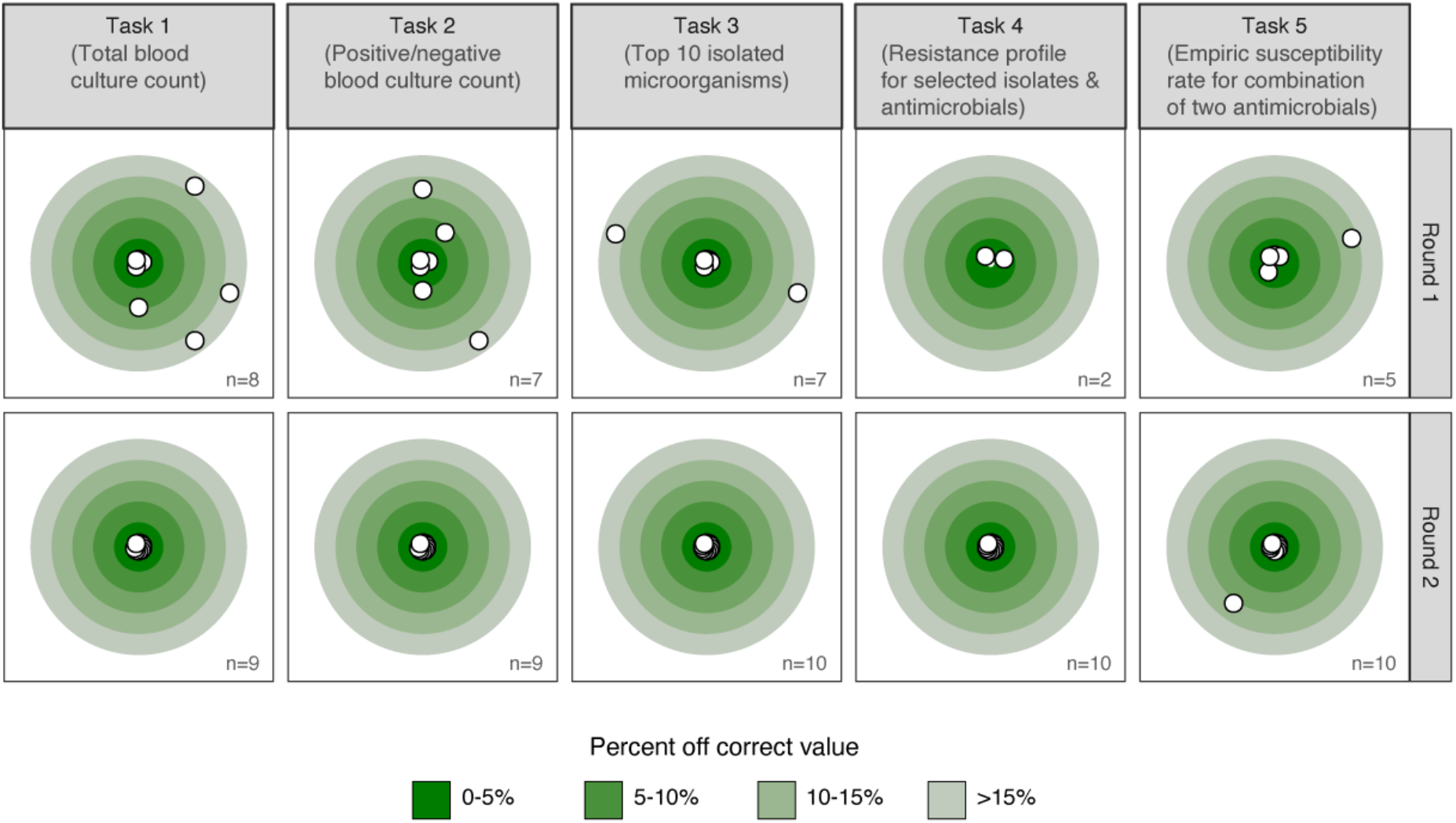
Deviation from the correct result by task and round in absolute percent from correct result. Only completed tasks (n) are shown.

### Efficiency

Overall, the mean time spent per round was significantly reduced from 93.7 (SD: 21.6) minutes to 22.4 (SD: 13.7) minutes (p < 0.001). Significant time reduction could be observed for tasks 2-5 (Figure 8). Analyses were further stratified to compare efficiency between participants that reported AMR data analysis as part of their job versus not part of the job. No significant time difference for completing all tasks could be found between the groups. However, in both groups the overall time for all tasks significantly decreased between the first and second round: on average by 70.7 minutes (p < 0.001) in the group reporting AMR data analysis as part of their job and by 72.1 minutes (p = 0.01) in the group not reporting AMR data analysis as part of their job.

**Figure 8.**
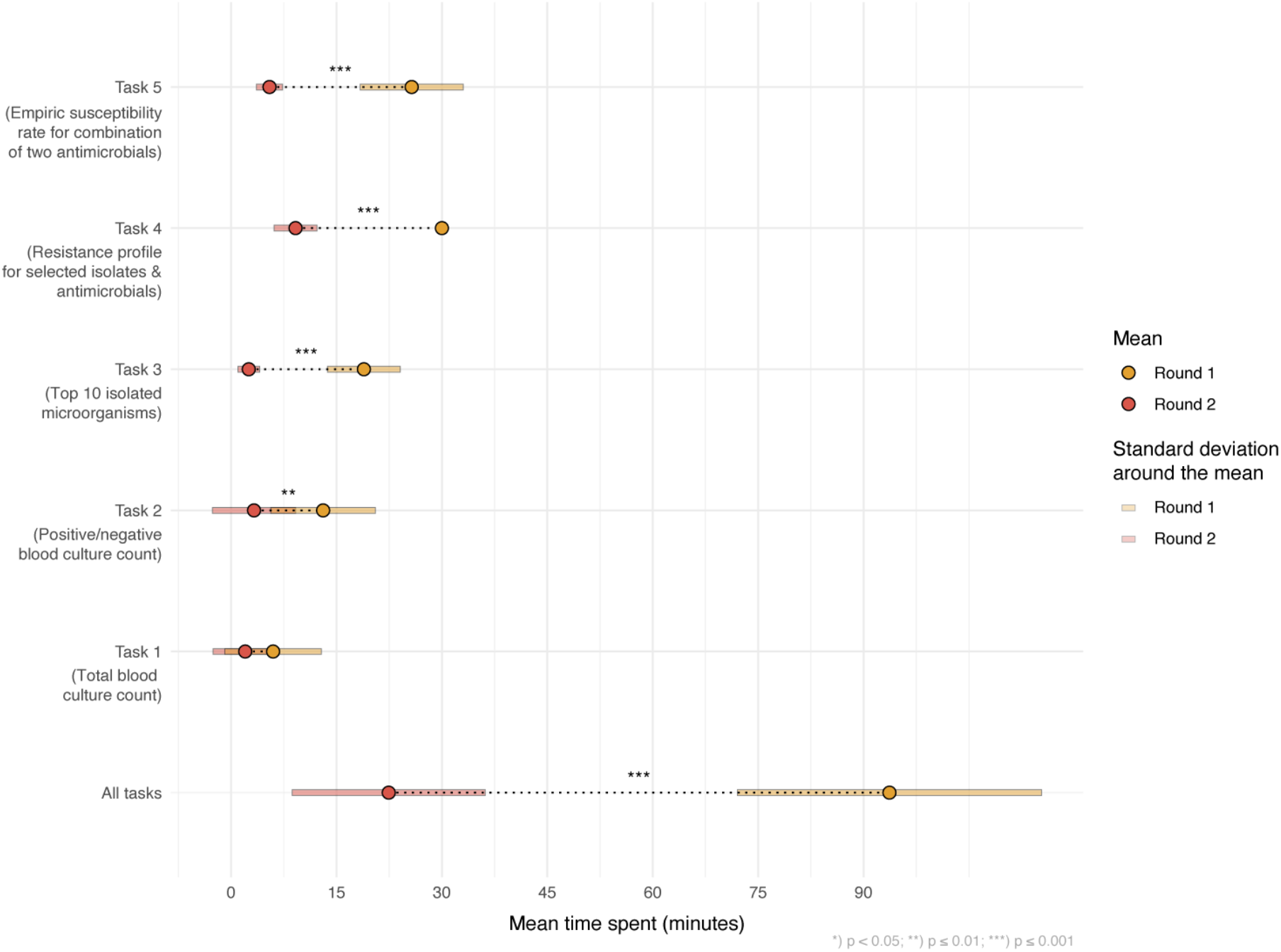
Mean time spent per task in minutes in each round (yellow = first round, red = second round). Statistical significance was tested using two-sided paired t-tests. All results were included irrespective of correctness of the results.

### Satisfaction

Participants rated the usability of the new AMR reporting tool using the system usability scale (SUS) which takes values from 0 to 100 (Appendix A2). This resulted in a median of 83.8 on the SUS.

## Discussion

This study demonstrates the effectiveness, efficiency, and accuracy of using open-source software tools to improve AMR data analysis and reporting. We applied our previously developed approach to AMR data analysis and reporting [7,8] in a clinical scenario and tested these tools with study participants (users) working in the field of AMR. Comparing traditional reporting tools with our newly developed reporting tools in a two-step process, we demonstrated the usability and validity of our approach. Based on a five item AMR data analysis and reporting task list and the provided AMR data, study participants reported significantly less time spent on creating an AMR data report (on average 93.7 minutes vs. 22.4 minutes; p < 0.01). Task completion increased significantly from 56% to 96%, which indicates that with traditional reporting approaches common questions around AMR are hard to answer in a limited time. The accuracy of the results greatly improved using the new AMR reporting approach, implicating that erroneous answers are more common when users rely on general non-AMR-specific traditional software solutions. The usability of our AMR reporting approach was rated with a median of 83.8 on the SUS. The SUS is widely used in usability assessments of software solutions. A systematic analysis of more than 1000 reported SUS scores for web-based applications across different fields has found a mean SUS score of 68.1 [18]. The results thus demonstrate a good usability of our approach.

The task list used in this study reflects standard AMR reporting tasks. More sophisticated tasks, such as the detection of multi-drug resistance according to (inter-)national guidelines were not included. However, these analyses are vital in any setting but restrained since the required guidelines are not included in traditional reporting and analysis tools (e.g., Microsoft Excel, SPSS, etc.). Notably, the underlying software used in this study [7] does provide methods to easily incorporate (inter)national guidelines such as the definitions for (multi-)drug resistance and country-specific (multi-)drug resistant organisms. The increase in task completion rate and accuracy of the results demonstrated that our tools empower specialists in the AMR field to generate reliable and valid AMR data reports. This is important as it enables detailed insights into the state of AMR on any level. These insights are often lacking. Our approach could fill this gap by democratising the ability for reliable and valid AMR data analysis and reporting.

This need is exemplified in the worrisome heterogeneity of the reporting results using traditional AMR reporting tools in the first round. Only 37.9% of the results in the first round were correct. Together with a task completion rate of 56%, this demonstrates that traditional tools are not suitable for AMR reporting. The inability of working in reproducible and transparent workflows further aggravates reporting with these traditional tools. All participants in the study should be able to produce standard AMR reports and 90% indicated that they worked with AMR data before. Sixty percent reported AMR data analysis to be part of their job, but no efficiency difference between groups were found. Our results show that AMR data analysis and reporting is challenging and can be highly error-prone. But an approach such as the one we developed can lead to correct results in a short time while being reproducible and transparent.

We chose an agile workflow which enabled us to integrate clinical feedback throughout the development process in this study. We can highly recommend this efficient approach for projects that need to bridge clinical requirements, statistical approaches, and software development. Our approach was inspired by others not in the AMR field that describe the use of reproducible open-source workflows in ecology [19]. We found that open-source software enables the transferability of methodological approaches across research fields. This transfer is a great example of the strength in the scientific community when working interdisciplinarily and sharing reliable and reproducible workflow.

This study is subject to limitations. Only ten participants were recruited for this study. Although low participant numbers are frequently observed in usability studies and reports show that only five participants suffice to study the usability of a new system, a larger sample size would be desirable [20–24]. In addition, other methods (e.g., ‘think aloud’ method) beyond the single use of the SUS for the evaluation of our approach would further improve insights in the usability but were not possible in the study setting [25]. Although the introduction of new AMR data and reporting tools made use of an already available approach, implementation still requires staff experienced in R. Reporting requirements also differ per setting and tailor-made solutions incorporating different requirements are needed.

The present study shows that answering common AMR-related questions is tremendously burdened for professionals working with data. However, answers to such questions are the requirement to enable hospital-wide monitoring of AMR levels. The monitoring, be it on the institutional, regional, or (inter-) national level, can lead to alteration of treatment policies. It is thus of utmost importance that reliable results of AMR data analyses are ensured to avoid imprecise and erroneous results that could potentially be harmful to patients. We show that traditional reporting tools and applications that are not equipped for conducting microbiology epidemiological analyses seem unfit for this task - even for the most basic AMR data analyses. To fill this gap, we have developed new tools for AMR data analysis and reporting. In this study, we demonstrated that these tools can be used for better AMR data analysis and reporting in less time.

## Data Availability

Data is available upon request

## Acknowledgement

We thank all study participants for their participation in this study and highly value their time spent on these tasks next to their clinical and other professional duties.

## Appendix

### A1) Task lists including correct results

**Table A1.**
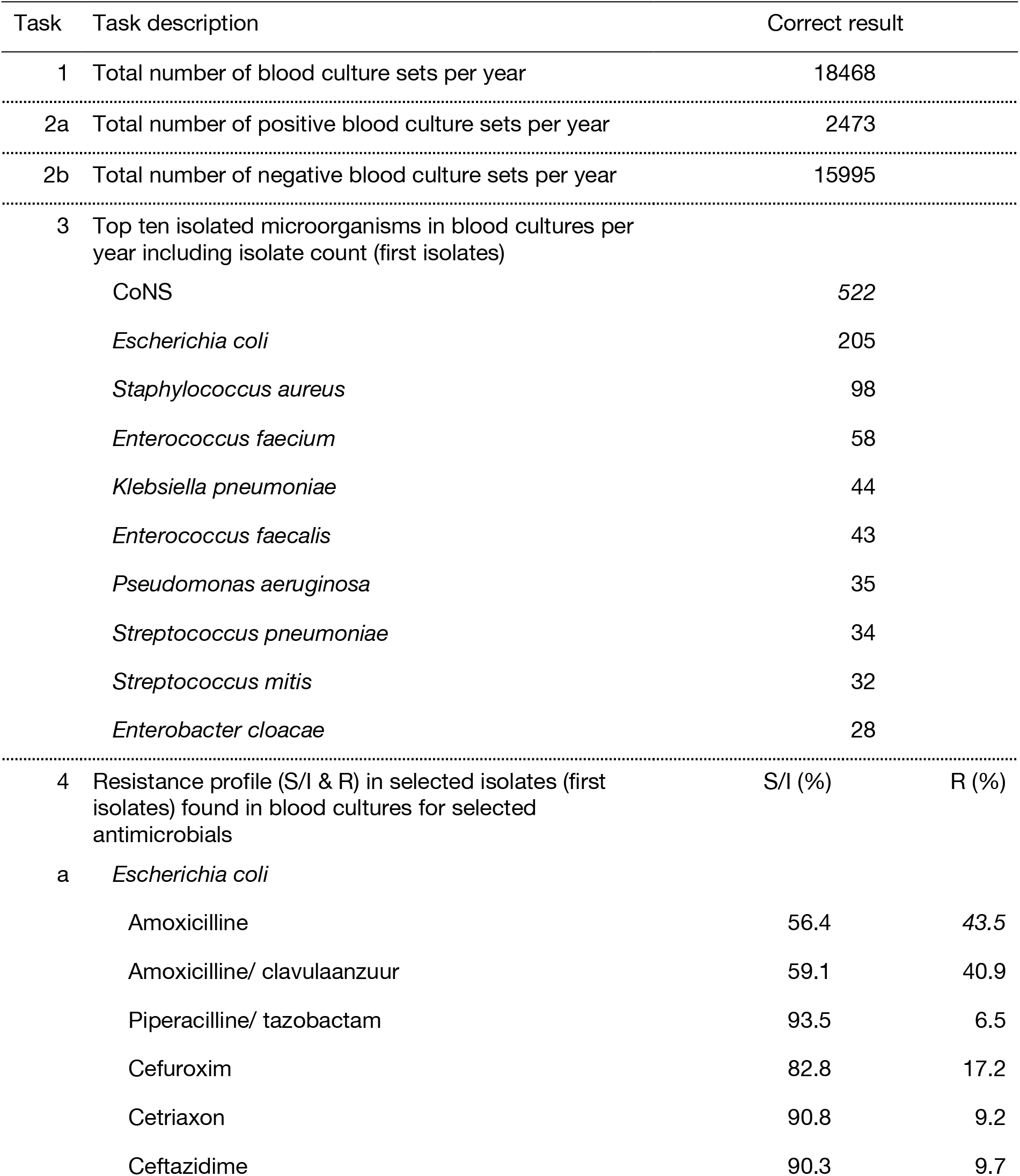
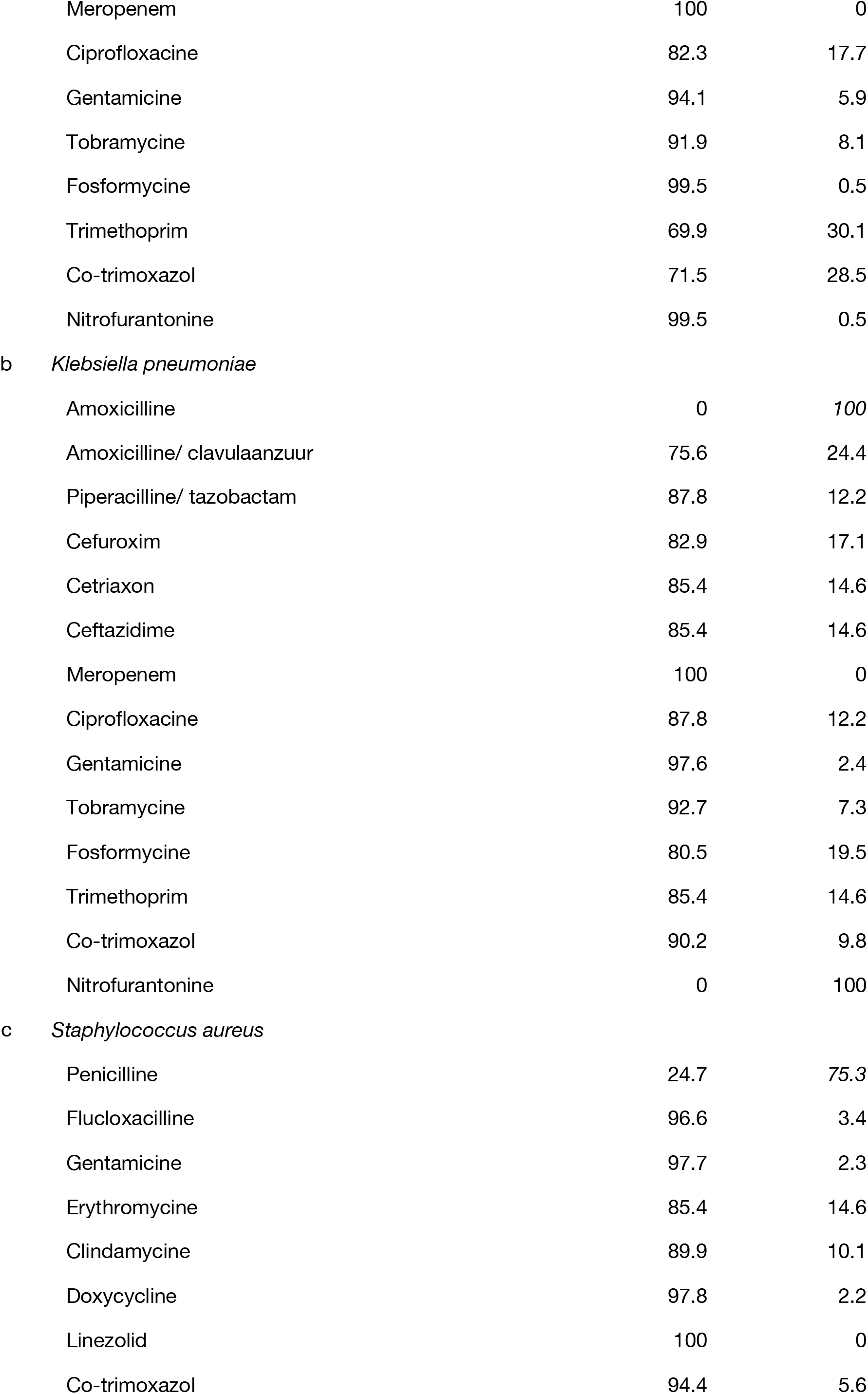
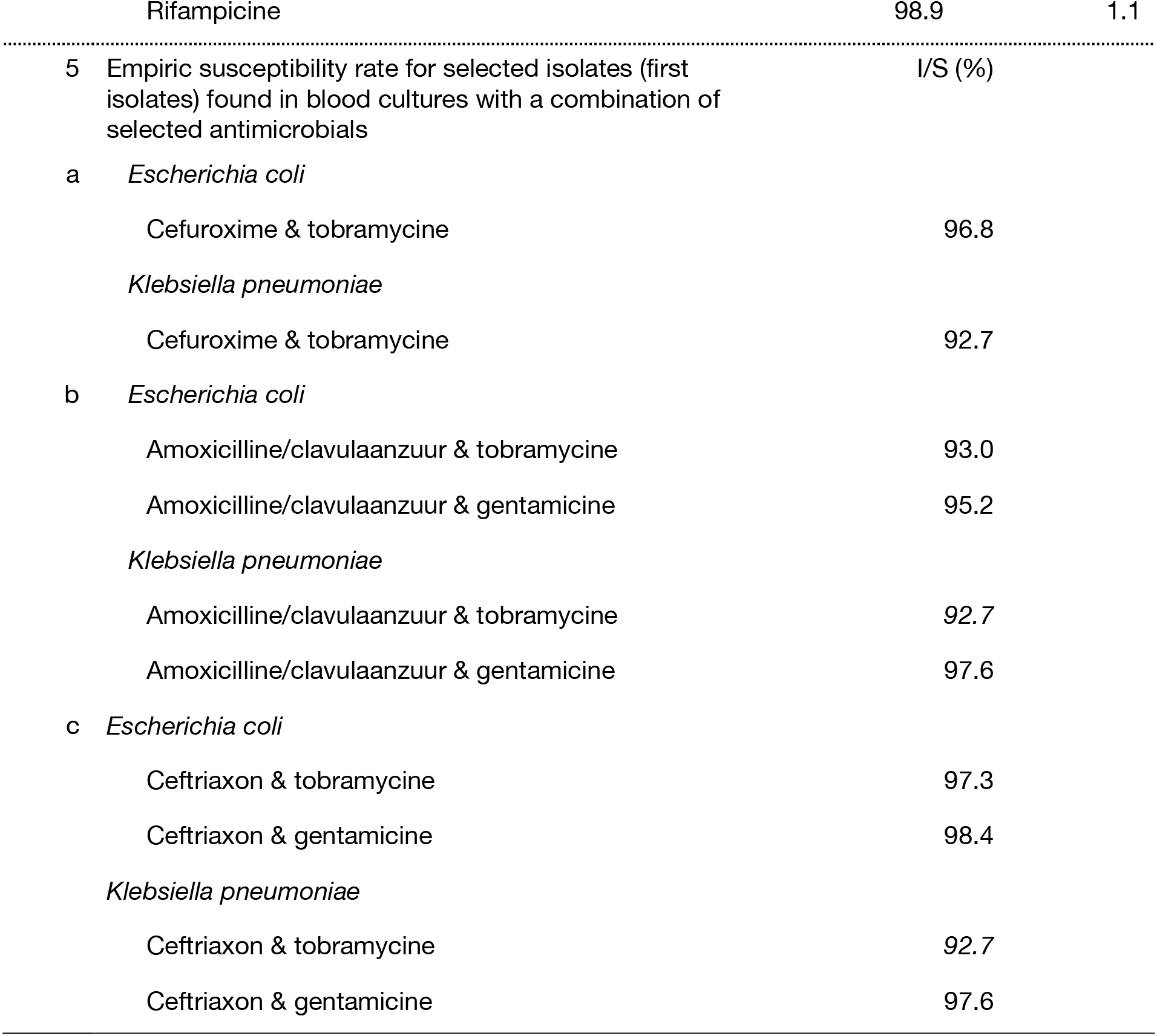
AMR data analysis and reporting tasks with correct results

### A2) System Usability Scale (SUS)

1. I think that I would like to use this system frequently.
2. I found the system unnecessarily complex.
3. I thought the system was easy to use
4. I think that I would need the support of a technical person to be able to use this system.
5. I found the various functions in this system were well integrated.
6. I thought there was too much inconsistency in this system.
7. I would imagine that most people would learn to use this system very quickly.
8. I found the system very cumbersome to use.
9. I felt very confident using the system.
10. I needed to learn a lot of things before I could get going with this system. (each item with levels: 1 = strongly disagree to 5 = strongly agree)

Scores for individual items are not meaningful on their own. To calculate the SUS score, the score contributions from each item must be summed. Each item’s score contribution ranges from 0 to 4. For items 1, 3, 5, 7, and 9 the score contribution is the scale position minus 1. For items 2, 4, 6, 8, and 10, the contribution is 5 minus the scale position. The sum of the scores is multiplied by 2.5 to obtain the SUS.

### A3) Task 3 sub-analysis

Task 3 asked participants to identify the ten most frequent species in the provided data set, while correcting for multiple occurrences of a species within a patient. Figure A2 illustrates the deviation from the correct result in the first round (traditional AMR reporting) per species. For this analysis also incomplete results were included (i.e., task not completed but some results provided).

**Figure A2.**
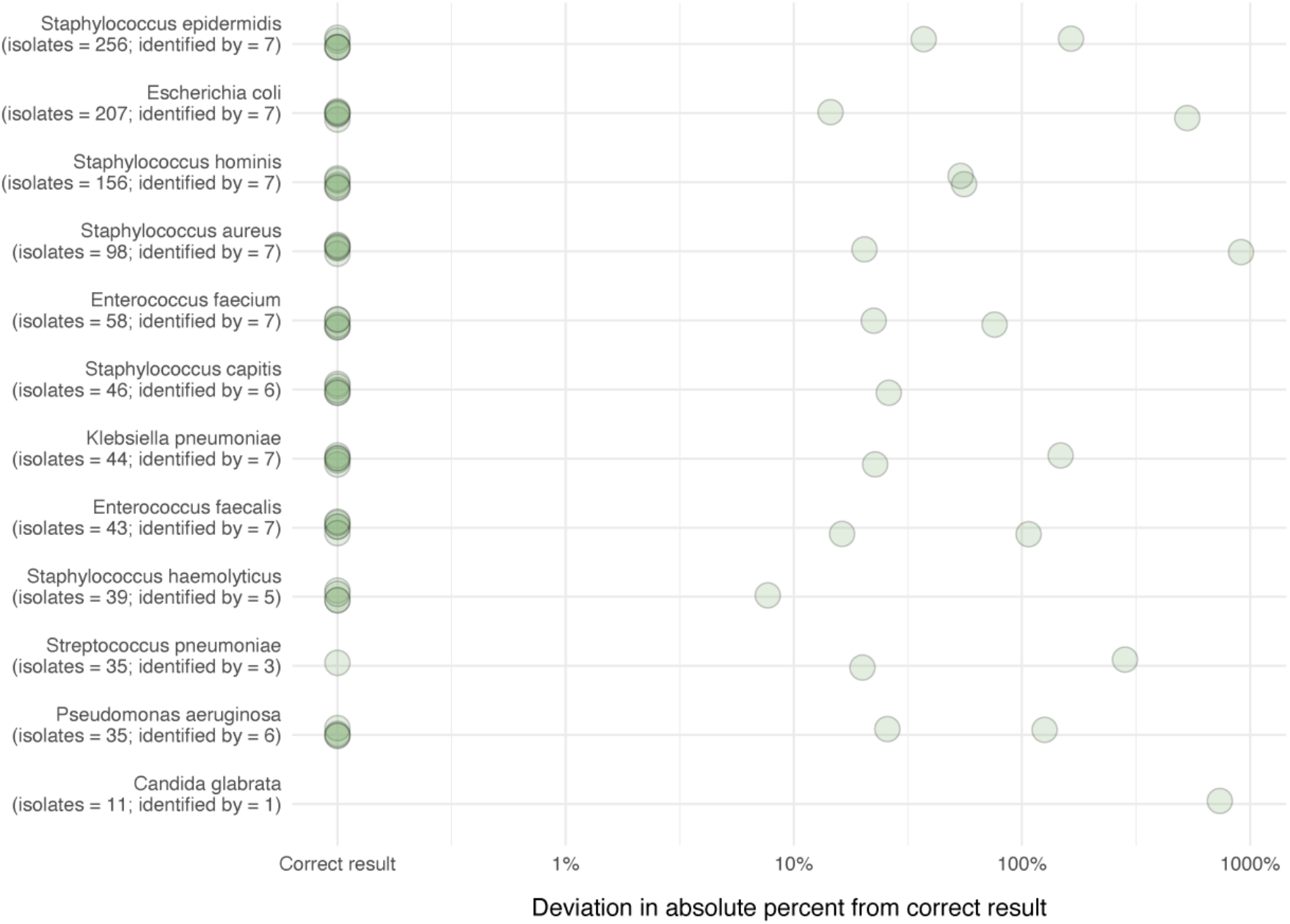
Results from task 3 in round 1. Deviation in absolute percent from the correct result per identified species. Also incomplete data from participants was used in this analysis (i.e., task not completed but some results given). The correct number per species is given in addition to the number provided answers.

